# Self-Burnout – A New Path to the End of COVID-19

**DOI:** 10.1101/2020.04.17.20069443

**Authors:** B Shayak, Richard H Rand

## Abstract

In this work we use mathematical modeling to describe a possible route to the end of COVID-19, which does not feature either vaccination or herd immunity. We call this route self-burnout. We consider a region with (*a*) no influx of corona cases from the outside, (*b*) extensive social distancing, though not necessarily a full lockdown, and (*c*) high testing capacity relative to the actual number of new cases per day. These conditions can make it possible for the region to initiate the endgame phase of epidemic management, wherein the disease is slowly made to burn itself out through a combination of social distancing, sanitization, contact tracing and preventive testing. The dynamics of the case trajectories in this regime are governed by a single-variable first order linear delay differential equation, whose stability criterion can be obtained analytically. Basis this criterion, we conclude that the social mobility restrictions should be such as to ensure that on the average, one person interacts closely (from the transmission viewpoint) with at most one other person over a 4-5 day period. If the endgame can be played out for a long enough time, we claim that the Coronavirus can eventually get completely contained without affecting a significant fraction of the region’s population. We present estimates of the duration for which the epidemic is expected to last, finding an interval of approximately 5-15 weeks after the self-burnout phase is initiated. South Korea, Austria, Australia, New Zealand and the states of Goa, Kerala and Odisha in India appear to be well on the way towards containing COVID by this method.

## §0 INTRODUCTION

In December 2019, the detection of a new Coronavirus disease did not raise any alarm anywhere. In the subsequent months however, the virus has spread to literally every country of the world. As we write this, there are more than 20,00,000 Coronavirus cases on record with 1,40,000 fatalities. The financial cost of the virus has been hardly less mind-boggling. Not since the time of the great Wars has there been a simultaneous stalling of all the world’s major economies, with almost our entire population being confined to their homes in actual or virtual quarantine. Unemployment ratios have reached unprecedented highs, and scarcity of food, in the most direct sense of the term, has become a serious concern everywhere. Nevertheless, amidst the various horror stories of death and devastation, the daily news reports have started providing some glimmers of hope. Italy, which was at one time the worst affected country, has by now exhibited a manifest reduction in the daily increments of cases and deaths. Spain, where the tally was increasing by 8000 cases/day at the peak, has now slowed down to about half that rate, beyond the purview of experimental error. A similar phenomenon has occurred in Germany. A marked reduction in rate can also be seen in Iran, where the maximal value of more than 3000 cases/day has since reduced to less than 2000 cases/day. The state of New York in USA has recorded three successive days of about 7500 fresh cases after a string of four consecutive five-figure daily increments – although three swallows are not proof of summer, there is some reason to believe that the worst is over for the Big Apple. With at least some regions entering a slowing down phase, we feel it shall not be premature on our part to start wondering as to how this scourge might actually end.

The end of a pandemic is something about which we have little prior knowledge. The two conventional ways in which pandemics end are (*a*) herd immunity and (*b*) vaccination. Herd immunity occurs when such a large fraction of the population is infected that the disease, so to speak, runs out of new unimmunized people to target. This is the way the influenza pandemics of 1957-8 and 1967-8 ended. In each case, about 1 lakh people were killed in USA alone. Nevertheless, they did not give rise to either the fear or the drastic restrictions which COVID-19 has generated. This was because they were low mortality diseases with death rates of a few percent of a percent – 1 lakh Americans were killed after almost the entire nation was infected. COVID-19 however is a high-mortality disease; if it is allowed to spread unchecked upto herd immunity, then the death toll in USA alone might run to eight figures. Vaccination is a fool- proof measure in theory – in practice, it takes 12-18 months to develop a certified usable vaccine. Then there are delays involved in creating and delivering the requisite numbers of doses to inoculate the entire world. Until that time, we cannot all remain in perpetual lockdown.

Every discussion of COVID-19 must also include SARS and MERS, which were also caused by coronaviruses and were extremely lethal. In both cases, the epidemics were contained with relatively little effort because the pathogens were quite difficult to transmit from one person to the next. The last time we’d had a highly transmissible and deadly pandemic had been the influenza of 1918-9. Unfortunately, back then, epidemiology did not exist as a scientific practice, and people did not keep very detailed records, so we have little knowledge of how that dreadful disease actually ended.

Media outlets are almost unanimous in their pessimistic proclamation [1-6] that COVID-19 can end only with vaccination or herd immunity. Social media tells a similar story. The scientific literature is less vehement but on the whole tends to agree. Kamikubo and Takahashi [7] and Moran et. al. [8] clearly indicate that the disease must progress upto herd immunity, while Das et. al. [9] and de Vlas and Coffeng [10] predict containment in two years (!) through intervention strategies. Mueller et. al. [11], Donsimoni et. al. [12] and La Scala et. al. [13,14] discuss various exit strategies from a lockdown without drawing any definite conclusion as to their effectivity or their expected time-frame. Kretzschmar et. al. [15] are clear that containment of the pandemic may be possible without resorting to herd immunity or vaccination; they do not give a timeline. Finally, at the optimistic end of the spectrum, Zhiglavski et. al. [16] predict the end of Coronavirus in England by August and Vattay [17] predicts the end in Italy by May. The former work however allows the disease to spread upto herd immunity while the latter is based on a logistic equation and appears overly simplified. In what follows, we shall describe a physically plausible scenario through which it is possible for the COVID-19 pandemic to be completely contained over a period of a few months without invoking herd immunity or vaccination. We call this the “self-burnout” route to the end of the pandemic.

## §1 MATHEMATICAL MODEL AND SOLUTION

We first demonstrate our strategy through a fantastical example. Let us say that the kingdom of Coronaland is suffering from corona cases when the King issues a decree that from now onwards every single person, well or unwell, must remain inside his/her home continuously until cleared to emerge. Because Coronaland is a fictional kingdom, this absolute lockdown does not generate a logistical problem of survival – sanitized robots, drones etc deliver food, medicines and other necessities to every single home. Those who had not been exposed to Coronavirus before the lockdown don’t contract the disease now. Those who had been exposed fall into three categories – the luckiest escape asymptomatically, the less lucky ones show symptoms but recover from it, and the unlucky ones die (inside their homes). As soon as someone dies, a robot sanitizes the body in a tankful of disinfectant. On the fourteenth day let us say after the lockdown, the last batch of the pre-lockdown exposures contracts the virus. On the twenty-first day, the last asymptomatic case among them recovers, on the twenty-fourth day the last unfortunate dies and on the twenty-eighth day the last symptomatic case overcomes the disease completely. On the twenty-ninth day, the King declares normal life back on in Coronaland.

When this resumption occurs, the virus does not exist any longer. Whatever the trajectory of cases before the 29-day lockdown, the number of new cases after the resumption will be identically zero. This happens because the virus can survive inside one particular person for only 28 days – within that time-frame, either the person’s immune system kills the virus or the person dies, which kills the virus also. The only way the virus can keep surviving beyond 28 days is if it can hop from person to person, which the lockdown prevents from happening. Starved of new targets for 29 days, the virus gets completely extinguished.

In Coronaland, it is possible to exorcise the pandemic through this simple strategy. In this land, it’s not. Nonetheless, we claim that in the real world also, we can feed the virus with new targets at such a slow rate that the disease eventually burns itself out. We now demonstrate this idea through mathematical modeling. Consider a localized region where there is no inflow of corona cases from the outside (we also ignore outflow although that will not ruin the endgame strategy for the region). We assume that the region, currently in a full lockdown, has reached a stage where the daily increments in cases have reduced to a significantly lower level than their peak values. This automatically implies that the regional authorities have the capacity to test (and treat) far more people than there are cases. The state of full lockdown can be lifted although it is vitally important to ensure that the increased horizontal separation minima of six feet between people and the absolute prohibition on large gatherings remain in effect. Even one mass transmission event can spell doom for the entire endgame strategy.

Multiple mathematical models of Coronavirus exist; we take as baseline the one which we ourselves developed three weeks back [18]. The technical advantage of this model is that it explicitly builds in the transmission of COVID by asymptomatic persons – those who remain asymptomatic throughout the infection period as well as those who develop symptoms after the latency period. Asymptomatic transmission is the primary difference between the present pandemic and the SARS and MERS outbreaks, so it is logical to use a model which accounts for this fact. The model in Reference [18] is Equation (7). It describes community spread during the bulk phase – we shall now modify it so as to account for the endgame phase. Figure 1 of that Reference forms the basis of the model derivation – it is absolutely general so we keep it for deriving the present model, and reproduce it below.

**Figure 1:**
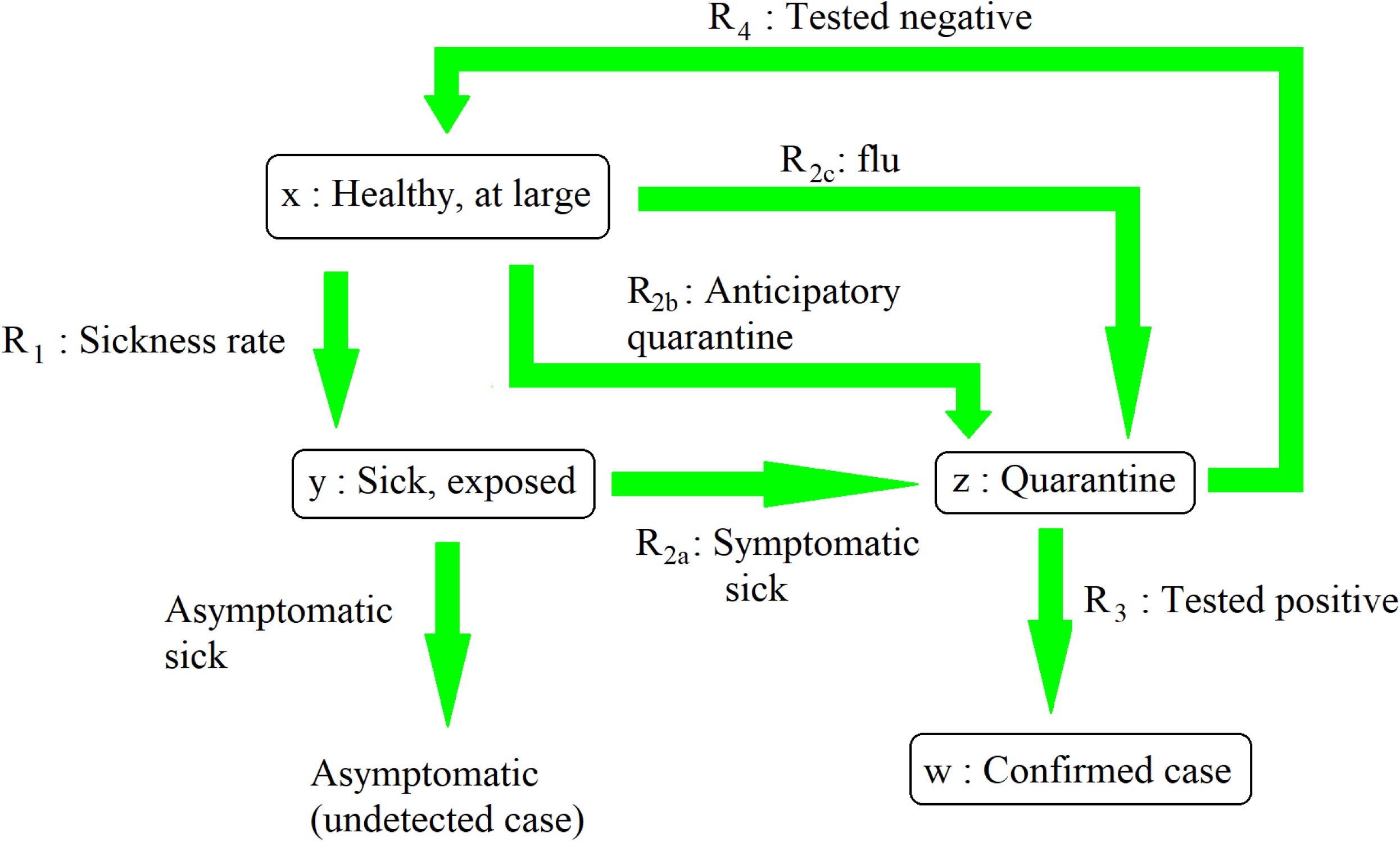
This Figure is taken from our own Ref. [18]. It is a flow diagram showing the various categories of people and the rates of transfer from one category to the other.

In this Figure, *x* is the number of healthy at-large people while *y* is the number of sick people who have had some exposure to society (i.e. they don’t fall sick in anticipatory quarantine). By definition, *y* is a cumulative counter – once a person falls sick, s/he remains a part of *y* for all subsequent time, even after s/he recovers or dies. We retain these definitions, as well as that of *R*_1_, the rate at which people transfer from *x* to *y*. In Ref. [18] we had taken *R*_1_ proportional to the number of healthy people and the number of sick people at large. This is an excellent assumption during a normal-life phase when every sick person (hereafter patient, even though we assume that s/he is at large only because s/he is asymptomatic) interacts with healthy people on the streets, in buses and trains, in restaurants and stores etc. The more the number of healthy people, the more the crowd through which the patient passes, and the greater the number whom s/he infects.

In a socially distanced phase however, this logic breaks down. If there is one patient on a bus, it does not really matter if there are two, five or ten other healthy people on the same bus, so long as all of them are at least six feet away from him/her and from each other, and are possibly wearing masks. Ideally, zero people get infected; practically there may be one or two “unlucky” transmission events which don’t really depend on the total number of passengers inside. If the bus is sanitized after each trip, the virus can be killed off and prevented from spreading any further. A similar phenomenon occurs in a grocery store where customers stand far apart from each other – ideally there is no person-to-person transmission. A more plausible transmission chain occurs if the patient handles an item which s/he does not buy. Since the virus remains on surfaces, it can get transmitted to the person or family who later buys the contaminated item. The transmission will likely get blocked however if the second customer quickly sanitizes every item which s/he buys. If the grocer him/her-self is a patient, s/he can spread the disease to all the customers. We can take care of the difference between grocer and customer through an averaging process (and after a point, these lumped-parameter models are heuristic anyway).

In the light of the above discussion, we decouple the *x*-dependence from *R*_1_ and claim that *R*_1_ depends on *y* alone through some constant of proportionality. An equivalent, though physically less insightful, interpretation of this is that due to small numbers of new cases, *x* remains almost constant throughout the endgame phase, so *k*_0_*x* in (7b) of Ref. [18] can be replaced by some new lumped parameter. We let *m*_0_ be the average rate at which one at-large patient infects healthy people i.e. we assume that, on the average, each patient infects in a time interval Δ*t* a number of people equal to *m*_0_Δ*t*. This average includes the grocer as well as the customer. The tacit assumption, inbuilt in all lumped parameter dynamical models, is that each patient infects other people continuously in time. To quantify the dependence of *R*_1_ on *y*, we need to go into the details of the contact tracing process.

In Ref. [18] we have treated the anticipatory quarantines arising from contact tracing (*R*_2*b*_) as a term disjoint from the people who fall sick in open society and report for quarantine (*R*_2*a*_). This is appropriate in a widespread community transmission situation where the overlaps between the anticipatory quarantines and the newly reporting cases will be negligible. In the endgame phase however, this will no longer hold true. With few new cases and high level of contact tracing, a significant fraction of the people whom one patient infects will end up being hunted down and quarantined before they fall sick.

Let Mr X be a patient who exhibits symptoms today and reports for quarantine. By the assumptions of the model, he has remained sick for the past *τ*_2_ days where *τ*_2_ is the latency period. During this time, he has infected other people continuously at a rate *m*_0_, so there are a total of *m*_0_*τ*_2_ new infections which have cropped up on Mr X’s account. As soon as he reports for quarantine, the authorities, we assume instantaneously, obtain his movements over the past *τ*_2_ days, searching frantically for potential patients. Let us say they track down and quarantine a fraction 1−*μ*_3_ of all the people whom Mr X has infected, where *μ*_3_ is a number between 0 and 1. (The similar quantities *μ*_1_ denote the fraction of asymptomatic patients relative to total patients, and *μ*_2_ was relevant only in Ref. [18] but has no role here; we let 1−*μ*_3_ rather than *μ*_3_ be the fraction of captures since that will simplify the final expressions a little.) Now, these captures will include people whom Mr X has infected right when he turned transmissible (i.e. *τ*_2_ days ago) and people whom he infected just prior to manifesting symptoms (i.e. right now). On the average, these contacts will have remained at large for a duration *τ*_2_/2.

As for the fraction *μ*_3_ of Mr X’s contacts who escaped the authorities’ notice, 1−*μ*_1_ turn symptomatic a time *τ*_2_ after sickness and head to quarantine on their own. Thus, they infect people continuously for the entire latency period *τ*_2_. Finally, the fraction *μ*_1_ of these escaped contacts is asymptomatic so they remain at large and transmit for the entire duration of the infection period *τ*_1_.

Putting all this together, we can say that patients belong to three classes – contact-traced, escaped symptomatic and escaped asymptomatic. The first class accounts for fraction 1−*μ*_3_ of all sick people and transmits for the duration *τ*_2_/2, the second class accounts for fraction (1−*μ*_1_)*μ*_3_ and transmits for the duration *τ*_2_, and the third class accounts for fraction *μ*_1_*μ*_3_ and transmits for the duration *τ*_1_. We now use the logic which we had already developed in Ref. [18]. Let us say a certain class of sick person *Y* transmits for a time duration *τ*. Then, new healthy people can only be infected by those *Y*’s who have fallen sick within the last *τ* days, and not the ones who have fallen sick earlier (recall that *y* and hence *Y* is a cumulative count – it includes every sick person including the one who fell ill right at the start of the outbreak). The number of such *Y*’s is their total count today minus their total count *τ* days ago, which is *Y* (*t*) − *Y* (*t*−*τ*). Since each such person transmits at constant rate *m*_0_, the contribution of class *Y* to *R*_1_ becomes *m*_0_ (*Y* (*t*) −*Y* (*t* − *τ*)). We now let *Y* denote each of the three classes (contact traced, escaped symptomatic and escaped asymptomatic) and add their contributions to *R*_1_, getting 

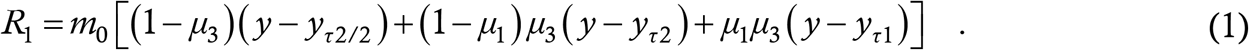

Here, subscripts indicate delay, as they do in Ref. [18]. Just as in there, d*y*/d*t* = *R*_1_. Simplifying, we get 

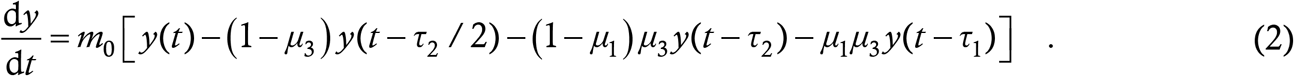

This is a single first order linear delay differential equation (DDE) governing the time evolution of the number of sick people.

Although (2) might appear like a drastic simplification relative to (7) of Ref. [18], it is in reality not all that different. Equation (7b) of Ref. [18] was nonlinear only through the *x* term which does not appear in a socially distanced endgame phase. Removing this dependence from (7b) of Ref. [18] and inserting the sizeable effect of anticipatory quarantine gives us (2). As regards *z*, that is of interest for estimating the total number of quarantine, treatment etc facilities required during the peak of the epidemic but it is not really useful during the endgame phase (we assume they are adequate). Finally, the number of cases *w* will be more or less the same as *y*, with retardations to account for latency and testing delay. Since the former is quite small, and the latter is negligible by our assumption of high testing capacity, *w* too is not of especial significance as an independent variable. We also note that the model of Ref. [18] was expressly designed for community transmission, and in all simulations there we assumed that the epidemic somehow ended after a 100-day period. Equation (2) here describes how this might happen. To summarize and emphasize the point once more, (2) is derived from the same underlying principles as (7) of Ref. [18], but it is NOT a simplified form of the latter which we have adopted for calculational purposes. Rather, it is as faithful a lumped-parameter model as we could come up with of the dynamics of corona cases during the controlled endgame phase, in which the authorities intend the epidemic to burn itself out.

When all the delays are zero, (2) becomes an ODE which has the solution *y* = const. We can see that *y* = const. is also a solution of the full DDE (2). So, if we try a solution to the DDE of the form e^*λt*^ where *λ* is an eigenvalue, then one of these eigenvalues will be zero. The stability of the constant solution will be governed by the other eigenvalues – if the real part of the largest eigenvalue is negative then a constant solution will be stable while if that real part is positive then a constant solution will be unstable. The former case corresponds to termination of the epidemic in time while the latter case corresponds to explosion of the epidemic in time. When the two delays are very small, we find that the solutions of (2) are stable, while they become unstable if the delays are increased. We now seek the condition for the stability transition.

The characteristic equation of (2) is, where we let *τ*_4_ = *τ*_2_/2, 

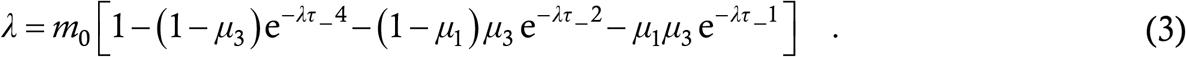

We can see that *λ* = 0 is always a solution. When the delays are identically zero, all the other eigenvalues have a real part of −∞ [19]; when the delays are increased, they start approaching the imaginary axis. The transition occurs when they actually breach this axis. There are two ways this can occur – a real root crosses the axis in a saddle node bifurcation or a pair of complex roots cross the axis in a Hopf. We first try the latter case. In this case, right at the transition point, there will be one pair of roots having the value ± j*ω* for some non-zero frequency *ω* [19]. Substituting this into (3) and equating the real and imaginary parts we get

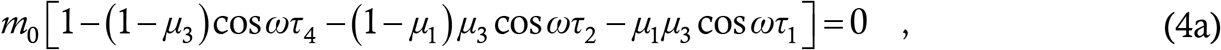

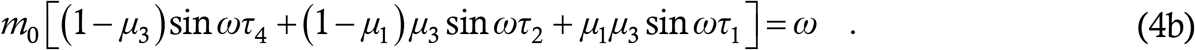

Now, (4a) has the structure 1− *a*C_4_ − *b*C_2_ − *c*C_1_ = 0 where C_1_ denotes cos *ωτ*_1_ etc and *a, b* and *c* stand for the three constants involved. This implies that *a*C_4_ + *b*C_2_ + *c*C_1_ = 1. But, *a,b,c* are positive and satisfy *a*+*b*+*c* = 1. Hence, if they are weighted by any kind of cosines, their sum will no longer will be 1, unless each of the cosines is itself 1. This must be the case in (4a). Immediately, each of the sines in (4b) becomes zero, and that equation reduces to *ω* = 0, which contradicts the starting premise of the case. Hence, (2) cannot undergo a Hopf bifuraction.

With Hopf ruled out, the transition to instability in (2) has to be through zero crossing of a single real root. At the instant of the transition, that will be a real root having the value zero. Since *λ* = 0 is already a root, at the transition it will become a repeated root. This means that (2) will have a solution proportional to *t* in addition to a constant solution [20]. The stability criterion comes out by forcing that *y* = *t* be a solution of (2); we immediately get

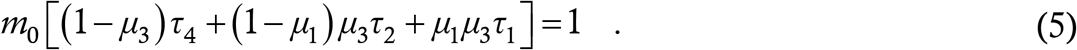

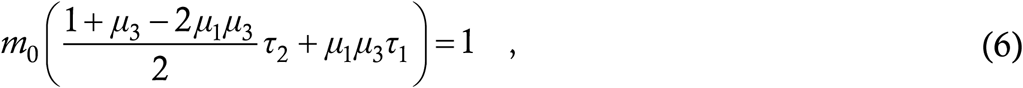

Recalling that *τ*_4_ was defined to be half of *τ*_2_, the criterion reduces to which is our final expression. The solutions of (2) are stable if the term on the above left hand side is less than 1 and unstable if this term is greater than 1. We have verified this criterion against numerical integration of (2), and in the next Section we shall discuss its implications.

Before launching that discussion, we present a plot of the transition curve in the *τ*_1_-*τ*_2_ plane for given values of the other parameters. We take the parameter values *μ*_1_ = 1/5 and *μ*_3_ = 1/2, and plot the transition curve (6) for *m*_0_ taking each of the four values 1/4, 1/3, 1/2 and 1.

The curve in each case is a straight line with the region to the left being stable and the region to the right being unstable.

## §2 IMPLICATIONS OF THE MODEL

The stability criterion (6) has a transparent interpretation. Recall that *m*_0_ is the basic rate at which each person transmits the infection, while the *τ*’s stand for the latency and the infection period. Equation (6) basically amounts to the condition that one sick person must not transmit the disease to more than one person on the average, during the entire time that s/he is transmissible and at large. This is of course a well-known criterion in terms of the reproduction number *R*_0_; what (6) does is to define it in a very precise manner in terms of social distancing measures (manifest through *m*_0_, more discussion later), latency period, asymptomatic transmission period, contact tracing activities etc. For example, if *μ*_3_ = 1 i.e. there is no contact tracing and *μ*_1_ = 0 i.e. there are no asymptomatic carriers either, then *τ*_2_*m*_0_ = 1 i.e. one patient should not transmit to more than one person during the entire latency period. The parameter *μ*_3_ is a measure of how many people escape contact tracing – the greater its value, the smaller must *m*_0_ be so as to meet the criterion. Similarly, *μ*_1_ is the fraction of asymptomatic carriers, who can transmit for a longer duration than the symptomatic ones – the greater this fraction, the smaller must *m*_0_ be again. These are basic physical plausibility criteria which our condition (6) satisfies.

In (6), *τ*_1_, *τ*_2_ and *μ*_1_ are parameters which are outside one’s control while *μ*_3_ lies with the regional authorities. The fact that higher contact tracing can permit greater social mobility is neither new nor profound. Of more significance are the measures which the authorities can take to limit *m*_0_ while maintaining a maximum possible social mobility, the latter being highly desirable – indeed, sometimes absolutely necessary – from the economic standpoint. The two factors affecting the average value of *m*_0_ are the rate at which close interactions (from the disease transmission viewpoint) occur between people and the probability that each such interaction actually does result in transmission. The first factor depends on the degree of restrictions on people’s mobility. For example, let us say that a patient has a 50 percent probability of infecting someone every time s/he goes out. In a full lockdown, s/he probably leaves home once every week, corresponding to a contribution to *m*_0_ of 1/14. But in a partial restriction situation, s/he will probably leave home once every alternate day, bringing *m*_0_ up to 1/4.

An important step towards reducing *m*_0_ consists in recognizing that some people have the potential to infect far more people per day, and hence contribute much more to the average, than others. Recall our example of the sick customer who infects possibly one person through a possibly tainted item and the sick grocer who infects a dozen customers by handling their goods with dirty hands. In an endgame phase, we expect the region to have a testing capacity significantly exceeding the actual numbers of tests being needed per day. The surplus capacity can be used to continuously (daily) test those people like the grocer, who can amount to major spreading risks. The moment such a person tests positive, s/he can immediately be placed into quarantine and out of harm’s way. The success of this measure is contingent on the test’s being fast – if it takes 5 days to process the results, then the measure becomes useless and the risk increases drastically. In regions where testing capacity is not that high, cluster testing [21] can be employed gainfully. For example, twenty grocers serving one neighbourhood can be tested in cluster every day, with individual testing being initiated only if a positive result is found. Note that we are asking for preventive testing only of the possible biggest spreaders, and not of the region’s entire population or a significant fraction thereof.

Measures such as sanitization, which breaks transmission chains perpetuated through material objects, and wearing masks, which shields people from virus in the event of a separation minima infringement, help to reduce the transmission probability associated with a close interaction, and thus also play a very significant role in reducing *m*_0_. Some recent studies [22,23] suggest that the transmissibility of the virus is reduced in environments having higher temperature and humidity. If this can be confirmed, then it will be useful to maintain high temperature and humidity inside potentially high-risk, climate-controlled environments like A/C buses and supermarkets.

The equation (2) and its stability condition (6) also tell us what kinds of relaxation of restrictions can be permitted during a post-lockdown self-burnout phase and what cannot. Lifting the six-feet separation minima is an obvious no, since (2) is contingent on the absence of unmitigated community transmission. With typical parameter values, the borderline *m*_0_ comes out to about 1/3 person per day (more on this later) i.e. one patient should spread the virus to maximum one person over three days. Since any person might be an asymptomatic patient, one person should on the average interact closely with at most one person every three days. This automatically rules out mass public gatherings since six-foot separation is not possible there, and a single positive case in such a gathering can transmit the disease like wildfire. Examples of mass gatherings which have led to corona disaster are the Tablighi Jamaat Islamic congregation in Nizamuddin, India and the Mardi Gras festival in New Orleans, USA.

In an academic institution, the faculty may be permitted to report for work since they sit in single offices. However, faculty meetings etc should be held via videoconferencing, and congregations in canteens should remain banned. Graduate students who are assigned multiply to a single office should not all report for work simultaneously. Per office, one student might perform on-campus work – the students can rotate on different days if the office is sanitized every day. Large research group meetings should remain on videoconference but a small in-person meeting such as 1-2 faculty interacting with 1-2 senior students or postdocs may be permitted once every 7-10 days. In-person classes should remain fully suspended but teaching staff may be allowed the use of a regularly sanitized lecture hall for delivering online classes instead of being forced to teach from home. Non-academic staff who process paperwork and interact with large numbers of people every day should continue working from home since they can amount to potential super-spreaders.

Long distance transportation should remain strictly on pause, since that can spread the disease among various regions and play havoc with the endgame strategy of each. Some of the excess vehicles and staff may be re-deployed for local transportation, where an increase in service can facilitate maintaining separation minima inside. The interiors should be sanitized frequently to prevent transmission chains via seats. On the social side of things, considerable mental stress is being generated in a region under lockdown by the unconditional prohibitions on any kind of socializing or outdoor activity. In a more relaxed endgame phase, a restaurant where every diner eats singly while sitting more than six feet away from each other should be permissible, with the safety factor increasing manifold if the tables and chairs are sanitized after every customer, and the food workers tested daily (possibly in cluster) for virus. Parties cannot be permitted. However, every person might be allowed to visit (or go out with) one friend once a week, which generates an average contribution to *m*_0_ of 1/7 if the person is a case. To accommodate this, restaurants might permit well-separated two-person tables say every Friday.

What makes a “region” ? In the Introduction we implied whole countries or states. Mathematically, this is acceptable since the model (2) does not depend on the region’s population and does not assume homogeneous mixing, as (7) of Ref. [18] does. Practically however, it is more realistic to define a region as smaller unit like a city, district or county. Not only will effective subdivisioning achieve better monitoring but it will also aid post-lockdown economic recovery since some regions shall achieve full containment before others. For example, Tompkins County (consisting primarily of the City of Ithaca) in New York State, where we are located, has registered 2 cases today, 3 yesterday and none at all on the preceding two days. This was possible on account of the early suspension of operations at our University, and the order to evacuate all undergraduate students ten days prior to the statewide lockdown. A significant fraction of Tompkins’ 118 cases arose from a single spreading event at a Chinese restaurant, where a waiter had the virus. This despite New York State’s being one of the world’s worst Coronavirus horror shows (this is the first time that the 5.5-hour distance from New York City has felt like a blessing).

If a region does achieve complete containment (no new cases for ∼14 days), then the most logical action is to hermetically seal its borders and throw it open internally. Normal life should be restarted inside the region while rigorously preventing transport of people to and from the outside, which is still contaminated. We recognize that material supplies from the outside might be essential to the region’s survival. To minimize risk, a consignment loaded into a truck outside the region should be kept inside the truck for 2-3 days, which is the estimated lifetime of the virus on surfaces [18,22]. Only thereafter should it be delivered to the virus-free region. Inside the latter region, unloading should be done entirely by residents of that region, with the driver remaining confined inside the truck at all times. Similarly, supply trucks travelling from a virus-free region to a contaminated one should undergo a change of driver and rigorous sanitization at the border.

If two regions declare successful containment of the virus, then transportation links can be opened up between them. Managing public transport is easy; the ease of controlling private transport will depend to a large extent on how much information can be obtained from a car’s license plate. For example, in India, a car’s home city is encoded into its number – in the state of Uttar Pradesh, the designations UP32, UP78, UP77 and UP70 denote the cities of Lucknow, Kanpur, Fatehpur and Allahabad respectively. In USA however, the city information is not written on its licence plate, so private cars will probably have to be kept off state highways and interstates for now.

In practice, *m*_0_ in (2) is a parameter which the authorities cannot calculate a priori, although they can possibly estimate it to some degree by conducting popular surveys of daily activities. All other parameters are known from virological studies or can easily be obtained from past data sets. To calculate the effective *m*_0_, the authorities can use the real time case data which they acquire. Here also, the absence of testing delay enters the picture – if delay is present then the case history is a faithless reproduction of *y*(*t*), as we have discussed in Ref. [18]. But when the recorded history is a more or less accurate reproduction of *y*, the effective value of *m*_0_ during say the past week can be found by entering the data from the week before into (2), and simulating. The simulation results (coming below) also show that, in the stable regime, if (2) is seeded with initial straight line data, the curve remains concave throughout until reaching the equilibrium. So, if at any time there is a tendency towards convexity, then social mobility should immediately be restricted further.

A question present in all our minds even if we dare not voice it is, how long will COVID-19 last ? We use our model to take a stab at this issue. For this we simulate (2) in Matlab, using the same numerical integration routine employed in our previous work. We take the parameter values *τ*_1_ = 7, *τ*_2_ = 3, *μ*_1_ = 1/5 and *μ*_2_ = 1/2 (the first three here are taken from Ref. [18], which includes a justification for their values). For this parameter combination, the critical value of *m*_0_ turns out to be 20/53 which is 0.3774. We seed the equation with a linear function *y* = 1000*t* (*t* in days) for the first 7 days (the maximum delay involved). We note that adding a constant *y* = 1000*t* +*C* does not change the subsequent case histories in any manner other than to add the constant *C* to the entire trajectory. 1000 cases/day is a realistic estimate for a post-peak hotspot region (Italy, Spain, New York State) which can consider lifting a total lockdown and initiating a controlled endgame phase. We define the epidemic to be over on the first day that there is less than one new case, and consider the endgame duration as the interval from the start of free evolution (excluding seeding) to the day that the epidemic is over. We also calculate the number of people infected during this time.

We find that the duration and severity of the epidemic in the endgame phase vary dramatically depending on how close *m*_0_ is to the critical value. At 50 percent of critical, the epidemic lasts only for 21 days, infecting another 1800 people in the process. At 80 percent, this increases to 59 days and 7250 people, at 90 percent, to 122 days and 16500 people, and at 95 percent to 248 days and 34500 people. The time trace of the epidemic in the 90 percent case is shown below.

What is a realistic value of this percentage ? In Ref. [18], it was South Korea which provided a data set for fitting, and once again it’s the same country which does the honours. For after a nearly linear profile of cases in the second half of March, South Korea is now clearly showing a downward-bending case trajectory indicative of an endgame phase. From 28 March to 04 April, the cases increased almost linearly from 9478 to 10156. The cases on the next few days until yesterday, as per Reference [24], are given in Table 1.

**Table 1:** Case history in South Korea between 05 April and 14 April 2020.

| Date | 05 | 06 | 07 | 08 | 09 | 10 | 11 | 12 | 13 | 14 |
| --- | --- | --- | --- | --- | --- | --- | --- | --- | --- | --- |
| Cases | 10237 | 10284 | 10331 | 10384 | 10423 | 10450 | 10480 | 10512 | 10537 | 10564 |

**Table 2:** Case history in Austria between 24 March and 24 April 2020.

| Date | 24/03 | 25 | 26 | 27 | 28 | 29 | 30 | 31 | 01/04 | 02 | 03 |
| --- | --- | --- | --- | --- | --- | --- | --- | --- | --- | --- | --- |
| Cases | 5243 | 5560 | 6847 | 7557 | 8233 | 8774 | 9541 | 10088 | 10711 | 11129 | 11521 |
| Date | 04 | 05 | 06 | 07 | 08 | 09 | 10 | 11 | 12 | 13 | 14 |
| Cases | 11766 | 11973 | 12297 | 12519 | 12942 | 13246 | 13560 | 13806 | 13945 | 14043 | 14234 |
| Date | 15 | 16 | 17 | 18 | 19 | 20 | 21 | 22 | 23 | 24 |  |
| Cases | 14350 | 14475 | 14595 | 14662 | 14710 | 14782 | 14833 | 14917 | 14932 | 15068 |  |

Using the parameter values mentioned before (for which the critical *m*_0_ = 20/53), we seed (2) with an exactly linear growth from 9478 to 10156 for 7 days and then pick the *m*_0_ which results in the best fit to the data of Table 1. We find that *m*_0_ equals 75 percent of the critical.

The overlap upto the 7^th^ day in the plot is insignificant since that is in the seeding period – the subsequent excellent match however is noteworthy. At this rate, the model predicts the epidemic to be fully contained 30 days after the seeing period i.e. on approximately 05 May. The total number of cases by then will be about 10680. Although we are not fool enough to bet money on these predictions, we do have confidence that the value of 75 percent is more or less accurate, and indicative. At this ratio, we find that the epidemic in the 1000 cases/day region considered in Fig. 2 burns itself out in 47 days, and infects 5450 people in the process.

**Figure 2:**
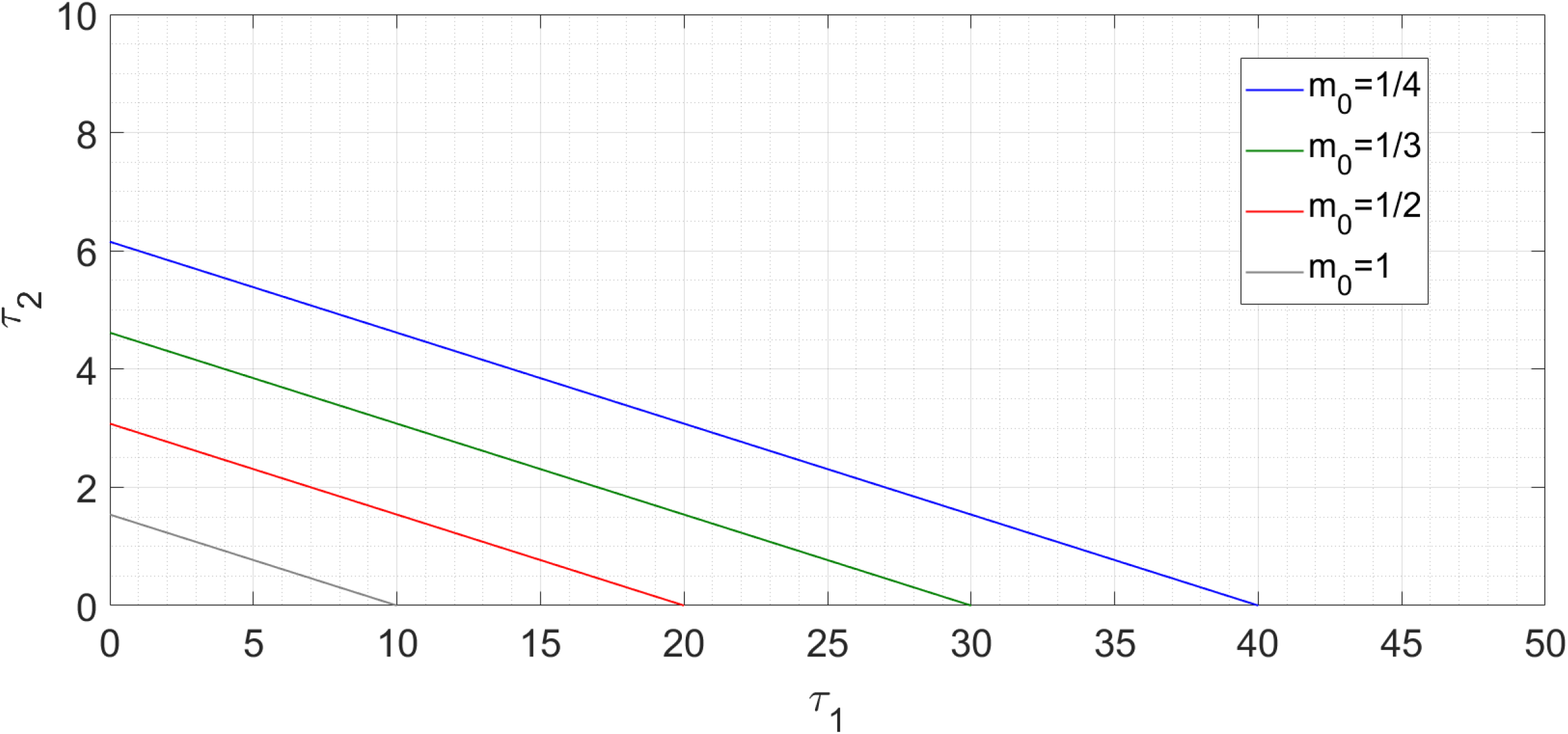
Stability transition curves for four values of m_0_. For each curve, the stable region is to the left and the unstable region to the right.

**Figure 3:**
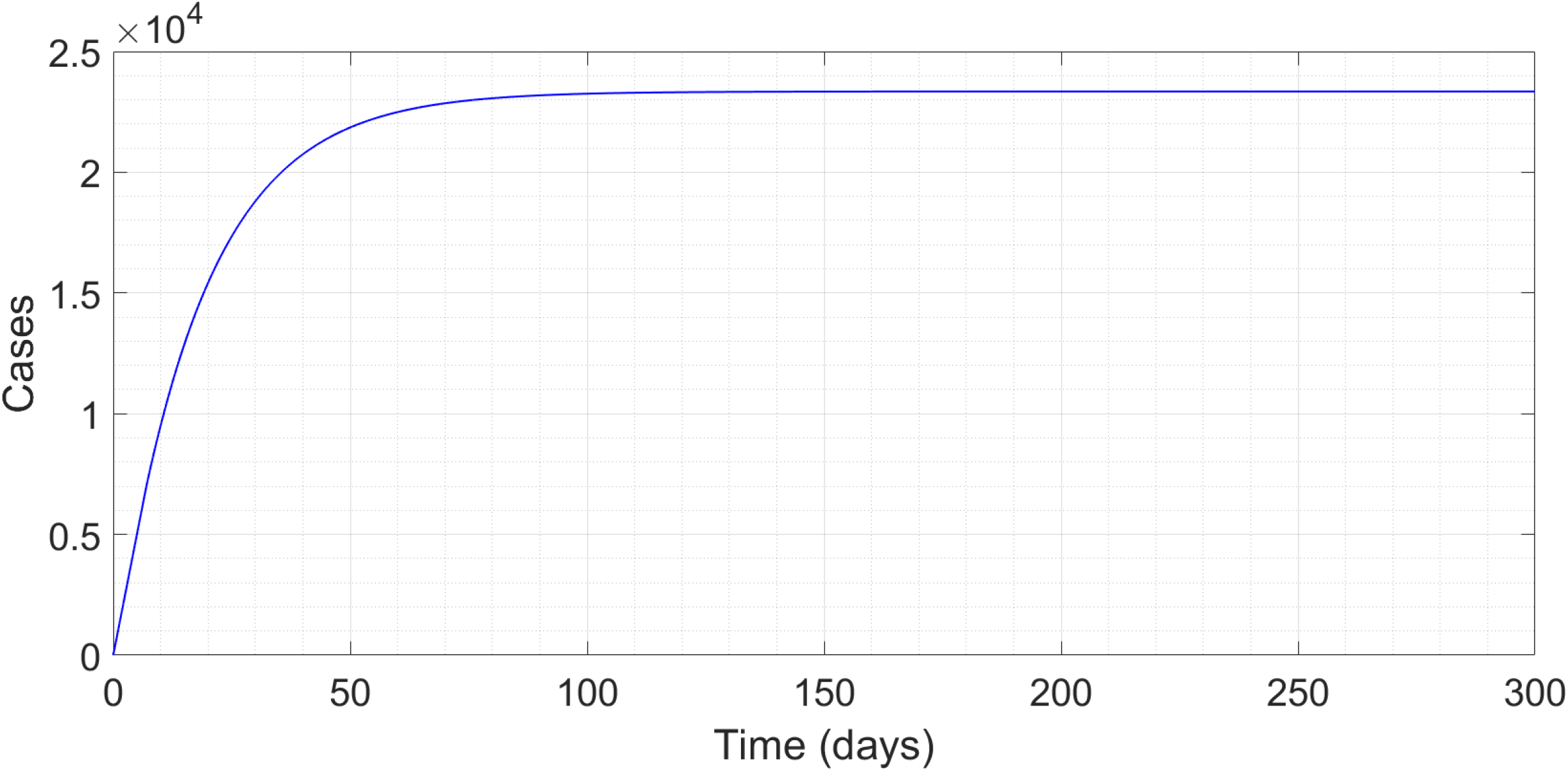
Time trace of epidemic with m_0_ being 90 percent of the critical.

**Figure 4:**
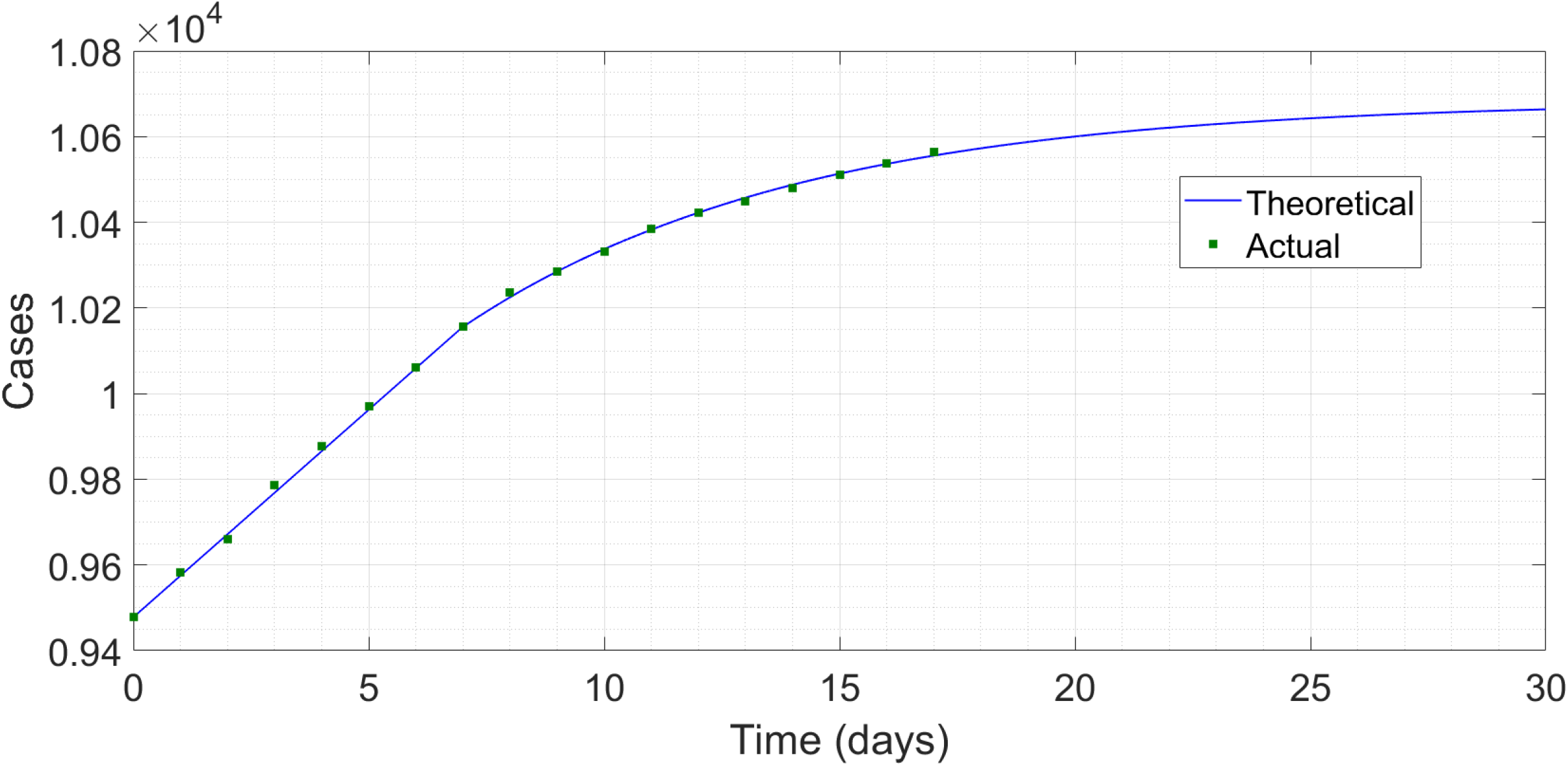
Comparison between predicted and actual case histories in South Korea.

We now investigate the sensitivity of the duration and severity to changes in parameter values. For this, we consider a parameter set *τ*_1_ = 10, *τ*_2_ = 4, *μ*_1_ = 1/5 and *μ*_3_ = 2/3. This is a far more adverse parameter choice than that of Fig. 2 and we have reason [25,26] to believe that these amount to a kind of worst case or worse-than-worst case scenario. In this case, the critical *m*_0_ comes out to 15/62, or just less than 1/4. Again we seed with 1000 cases per day for 10 days (the maximum delay). This time, at 50 percent of critical *m*_0_, we find 2800 more cases over 32 days, at 80 percent, 11,200 more cases over 91 days, and at 90 percent, 25,200 more cases over 189 days. Both the durations as well as the case counts are about 50 percent higher than in the previously simulated scenario – an acceptable level of variation.

Note that the time scale involved in self-burnout is one order of magnitude less than the time to termination of the outbreak through vaccination, and that the total number of cases is nowhere near that required to achieve herd immunity (tens of million cases per European country or American state). In reality, there will be testing errors, quarantine violations, illegal parties etc which will cause some disparity with our predictions, but we do not expect the epidemic duration or the case counts to increase by orders of magnitude. Also, towards the very end of the outbreak, with single-digit or teens of cases remaining in the region, a lot of probabilistic stuff will enter the picture and our model, which, like any lumped parameter study, is based on averages, will break down completely. These effects might easily add 2-3 weeks and dozens or even hundreds of cases to our estimates.

While a lot of our discussion mentions a post-lockdown scenario, (2) is also applicable to a hotspot which is currently under lockdown and has passed the peak. In lockdown, *m*_0_ will be extremely low, and for high case rates, contact tracing will be ineffective so *μ*_3_ will be high. While fitting near-peak regional data, we must also account for the delay in testing if that is present, as in Ref. [18]. A sample region can also be one where a definite peak has been avoided through early lockdown and then the endgame is being initiated with an increase in the testing capacity. Ithaca, which we have discussed previously, falls into this category. The state of Kerala in India [27] is likely in this regime and we hope that Nayi Dilli and Mumbai may also enter this category within the next couple of weeks.

## §3 CONCLUSION

In this Article, we have demonstrated how self-burnout – continuous social distancing combined with extensive testing and contact tracing over a period of weeks – can result in complete containment of the Coronavirus pandemic at a regional level without resorting to herd immunity and in a time frame significantly smaller than what the development of a vaccine would undertake. Multiple such regions can eventually make up a country, and finally the whole world. While restrictions on social mobility are likely to remain in place for a while, yet with some luck we are probably not looking at house arrest all the way upto 2022. The greater the restraint we practise now, the faster are we going to have this evil virus behind us. Our analysis also identifies the kinds of relaxation which can be permitted during the endgame phase, and the kinds which definitely cannot.

Another factor which probably works in our favour is that summer is just arriving or has already arrived in most of the affected regions. As already mentioned, there is evidence to indicate that the transmissibility of the virus is reduced in summer. The initial outbreak in Australia and New Zealand, which were in summer when the virus first arrived, has remained quite muted. In India and Singapore also, the trajectory so far has been flatter than in similar places located in colder climates. Reduced transmissibility means that for the same social restrictions, *m*_0_ will be lower. As we saw, even a small reduction in *m*_0_ can result in a drastic shortening of the time to the end of the outbreak.

Although we would not like to spawn false hope, we do think that there is a reasonable chance that COVID- 19 might really get contained within the end of summer. Of course, this will be contingent on the authorities’ doing a continuously competent job of epidemic management and everyone’s abiding by the restrictions that are laid down. Mistakes during the endgame, over-hasty opening of borders and similar actions can result in a virulent second wave being released and the good work of several past months being undone. However, with steady leadership from the scientific community and local authorities being supplemented by smiling-faced cooperation from all of us, it is not an absolutely fantastic possibility that by autumn, COVID-19 will have become history in most of the globe. In conclusion, the ultimate fate of COVID-19 rests on an Authority who is neither a scientist nor a doctor nor a politician – after describing one possible outcome which that supreme Authority might have in store for us, let us offer a prayer that this may indeed be true.

## Data Availability

All data sets used in this manuscript are publicly available.

## ACKNOWLEDGEMENT

SHAYAK would like to thank MOHIT SHARMA of Weill Cornell Medicine for valuable discussion.

## APPENDIX

This dates from Saturday, 25 April 2020. Here we incorporate some developments which have taken place over the past ten days. The rates in Italy, Iran and Germany have continued to slow down while Spain has seen an uptick in cases. New York State has passed its peak although the case rates remain high and a definite decline is yet to be seen. Mumbai, contrary to our hope of entering a burnout phase, has accelerated constantly to become India’s worst affected region while Dilli remains in a regime of steady growth.

On the brighter side of things however, there are several regions, both in India and abroad, which are definitely in a self-burnout phase. Goa, despite testing quite aggressively, has not found a single new case over the last 20 days [28], which almost certainly indicates that the outbreak has ended. Kerala and Odisha have brought down the daily increments to single digits. South Korea reported 50 additional cases shortly after we wrote our original draft – these were patients who had tested negative once but were found positive on a re-test – and the total now is 10708. Nevertheless, the overall decreasing trend is continuing, with the recent daily increments down to teens or single digits. Austria, Australia and New Zealand have also entered sustained phases of decline, which indicate self-burnout in progress. The raw data [23] from Austria (MOZART’s homeland, not the Southern Hemisphere country) has the least fluctuation so we have attempted a fit of that one. The data set is below.

We can see an approximately linear region upto 01 April, and a downward-bending region thereafter. So we use the 7-day interval 25 March to 01 April as the seeding period, after noting that by comparison with 24 and 26 March, the count for 25 March itself appears way too low. We correct this one-day error by fitting the next few days to a linear profile, finding a good fit for assumed 6250 cases on 25 March. We now feed this linear initial data into (2), just as we did for South Korea, using the same values for all parameters other than *m*_0_. This time we find the best fit for the subsequent evolution at *m*_0_ equal to 79.5 percent of the critical (compare with Korea’s 75 percent). The results are below.

**Figure A1:**
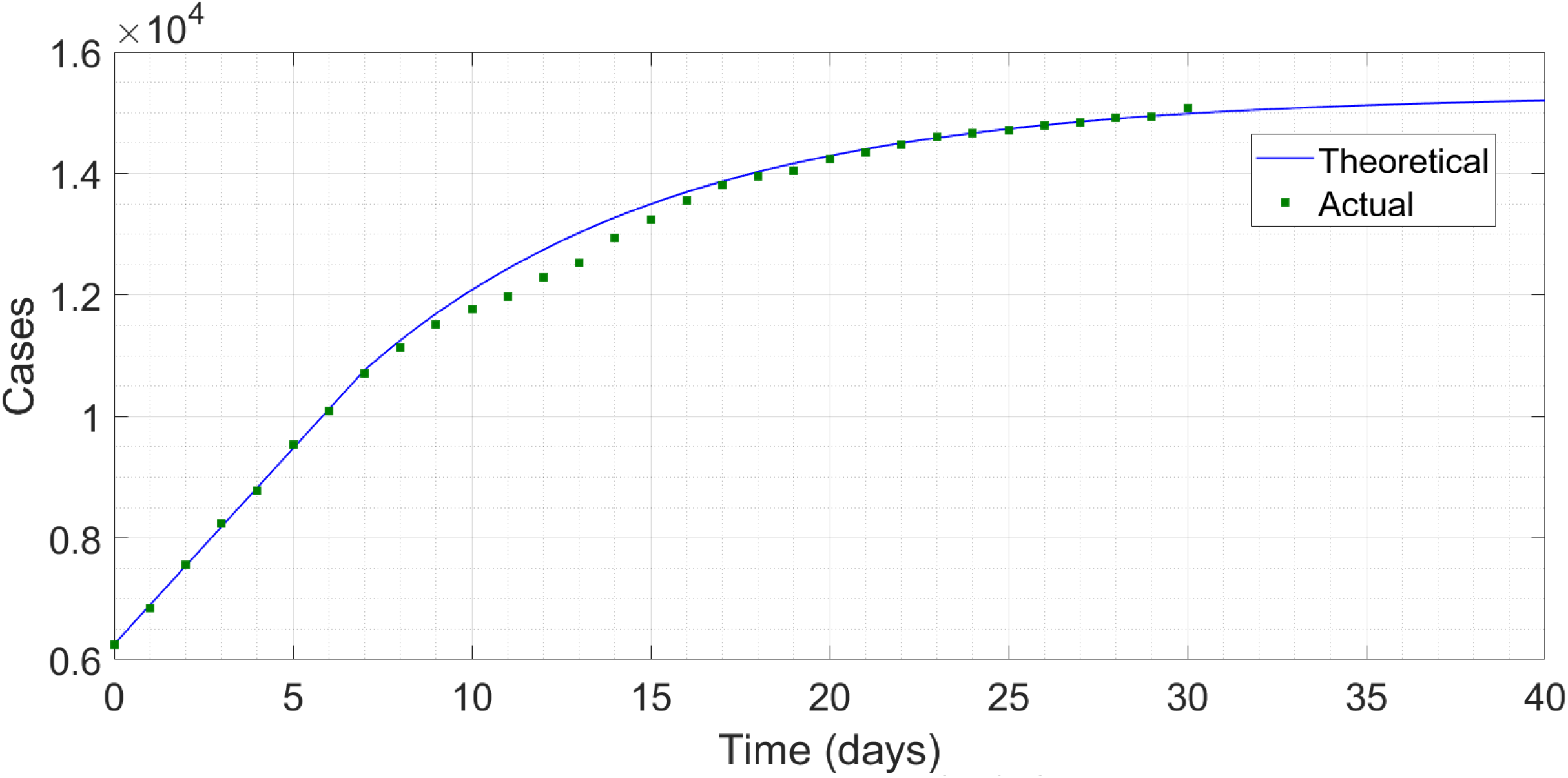
Comparison between predicted and actual case histories for Austria.

Once again, the first seven days correspond to the seeding period. Over the next eight days, the actual trajectory is lower than predicted but that might be a reporting error (the convexity in the actual data is implausible). This second quantitative agreement convinces us that self-burnout, as we have described it, is indeed working in multiple regions. So far, these regions have been ones where the initial impact of the disease was low. In a worse-hit region, the strategy takes longer to work but by no means fails, and we hope that the current hotspots will also use it to clamber out of this terrible pandemic.

